# The incidence and mortality of COVID-19 related TB disease in Sub-Saharan Africa: A systematic review and meta-analysis

**DOI:** 10.1101/2022.01.11.22269065

**Authors:** Jacques L Tamuzi, Gomer Lulendo, Patrick Mbuesse, Peter S. Nyasulu

**Affiliations:** Division of Epidemiology and Biostatistics, Department of Global Health, Faculty of Medicine and Health Sciences, Stellenbosch University, Cape Town, South Africa; Africa Centre for HIVAIDS management, Stellenbosch University, Cape Town, South Africa; Division of Epidemiology and Biostatistics, School of Public Health, Faculty of Health Sciences, University of the Witwatersrand, Johannesburg, South Africa

**Author notes:** **Corresponding Author:** Jacques L. Tamuzi Division of Epidemiology, Faculty of Medicine and Health Sciences, Stellenbosch University, Cape Town, South Africa. Emails: < >.

**Keywords:** COVID-19, PTB, Incidence rate, Case fatality rate, sub-Saharan Africa

## Abstract

**Background:** Coronavirus disease 2019 (COVID-19) is also associated with other co-morbidities in people who have previously or currently have pulmonary tuberculosis (PTB). PTB is a risk factor for COVID-19, both in terms of severity and mortality, regardless of HIV status. However, there is less information available on COVID-19 and PTB in terms of incidence and mortality rates in Sub-Saharan Africa (SSA), a high-burden TB region. This systematic review provided a data synthesis of available evidence on COVID-19/PTB incidence and case fatality rates, as well as mortality rates found in clinical and post-mortem COVID-19/PTB diagnostics in SSA.

**Methods:** We conducted an electronic search in the PubMed, Medline, Google Scholar, Medrxix, and COVID-19 Global literature on coronavirus disease databases for studies involving COVID-19 and PTB in Sub-Saharan Africa. The primary outcomes were the incidence proportion of people with COVID-19 who had current or previous PTB, as well as the case fatality rate associated with COVID-19/PTB. Based on methodological similarities in the included random effect model studies, the combination method was developed using Stata version 16 and Prometa 3 software. We also performed sensitivity analysis and meta-regression.

**Results:** From the 548 references extracted by the literature search, 25 studies were selected and included in the meta-analysis with a total of 191, 250 COVID-19 infected patients and 11, 480 COVID-19 deaths. The pooled COVID-19/PTB incidence was 3% [2%-5%] and a case fatality rate of 13% [4%-23%]. The pooled estimates for case fatality rate among COVID-19/PTB were 7% [1%-12%] for clinical PTB diagnostic and 25% [3%-47%] for post-mortem PTB diagnostic. Previous TB had the highest incidence and fatality rates with 46 [19-73] per 1, 000 population and 8% [3%-19%], respectively. Meta-regression model including the effect sizes and cumulative COVID-19 cases (P= 0.032), HIV prevalence (P= 0.041), and TB incidence (P= 0.002) to explain high heterogeneity between studies.

**Conclusion:** To summarize, the incidence of TB associated with COVID-19 is higher in SSA, as are the case fatality rates, when compared to the rest of the world. However, because the post-mortem TB diagnostic was higher, COVID-19 associated with TB may have been underreported in studies conducted in SSA. To confirm COVID-19/TB incidence and case fatality rates in SSA, large-scale cohort studies that adequately clear tools on previous and/or current TB diagnostic tools are required.

**Review registration:** PROSPERO (CRD42021233387)

## 1. Background

The coronavirus disease 2019 (COVID-19) pandemic has caused significant morbidity and mortality all over the world, with total confirmed cases more than 634 million globally and 6 million deaths in 25 November 2022 [1]. In the African region, more than 9 million confirmed cases are already recorded, among them, 174, 858 deaths on 25 November 2022[1]. Though it is reported that the African region showed a decreasing trend in the number of deaths over the past several weeks compared to other World Health Organization (WHO) regions [1]. The age groups at highest risk of severe COVID-19 disease and death (those >60 years old) [2-4] may be proportionately less in many sub-Saharan Africa (SSA) countries than in other parts of the world [4]. In contradistinction to potential health vulnerabilities, sub-Saharan African countries could be “protected” from COVID-19 mortality by an age structure differing significantly from countries where morbidity and mortality have been particularly high such as Italy, Spain, the United States, and in Hubei Province in China [5-7].

On the other hand, COVID-19 is associated with other co-morbidities in patients with previous or current pulmonary tuberculosis (PTB) [8]. A recent review found that PTB is a risk factor for COVID-19 regardless of HIV status, both in terms of severity and mortality [8]. Geographically, most people who developed tuberculosis (TB) live in South-East Asia (44%), Africa (25%), and the Western Pacific (18%) [9]. It is estimated that low- and middle-income countries, including SSA, account for 94% of all TB infections and deaths [10]. An observational study found that people with latent or active tuberculosis were more vulnerable to infection with the severe acute respiratory syndrome coronavirus 2 (SARS-CoV-2) [11]. This study found infection with *Mycobacterium tuberculosis* to be a more common co-morbidity for COVID-19 (36%) [11]. They also found *Mycobacterium tuberculosis* co-infection to be linked to more severe COVID-19 and more rapid progression [12]. Another study conducted in Zambia revealed that 31.4% of post-mortem PTB diagnoses in COVID-19 deaths [13]. In countries where PTB risk factors for mortality are highly prevalent among young individuals (poverty, overcrowding, diabetes mellitus, smoking, alcohol and substance abuse, HIV co-infection, among others), particularly in the presence of drug resistance and difficult access to diagnosis (delayed diagnosis) [14, 15], COVID-19 incidence and mortality associated with PTB may be hypothesized as significant. However, COVID-19 associated with previous, current, or both PTB has been lower in high-burden TB countries. Both active and previous history of PTB may play a deleterious synergism SARS-CoV-2, increasing then the risk of COVID-19 associated mortality, and for patients with PTB may increase the severity of COVID-19 and of death due to chronic lung disease and immunosuppression [8]. This may contribute to higher-than-expected mortality in high PTB-burden regions such as SSA.

Therefore, there is less information available on COVID-19 associated with PTB from the point of view of incidence and fatality rates in SSA. This systematic review served to provide data synthesis of available evidence on COVID-19/TB incidence and case fatality rates, and case fatality rates found in clinical and post-mortem COVID-19/TB diagnostics in SSA.

## 2. Results

### 2.1. Search results

From the 548 references extracted by the literature search (**Figure 1**), 64 articles were analysed, among which 33 were excluded with reasons, and 25 studies were selected and included in the meta-analysis and six studies were excluded as they were study protocols. We included a total of 191, 250 COVID-19 infected patients and 11, 480 COVID-19 deaths (**Supplemental material Table 1**).

**Figure 1:**
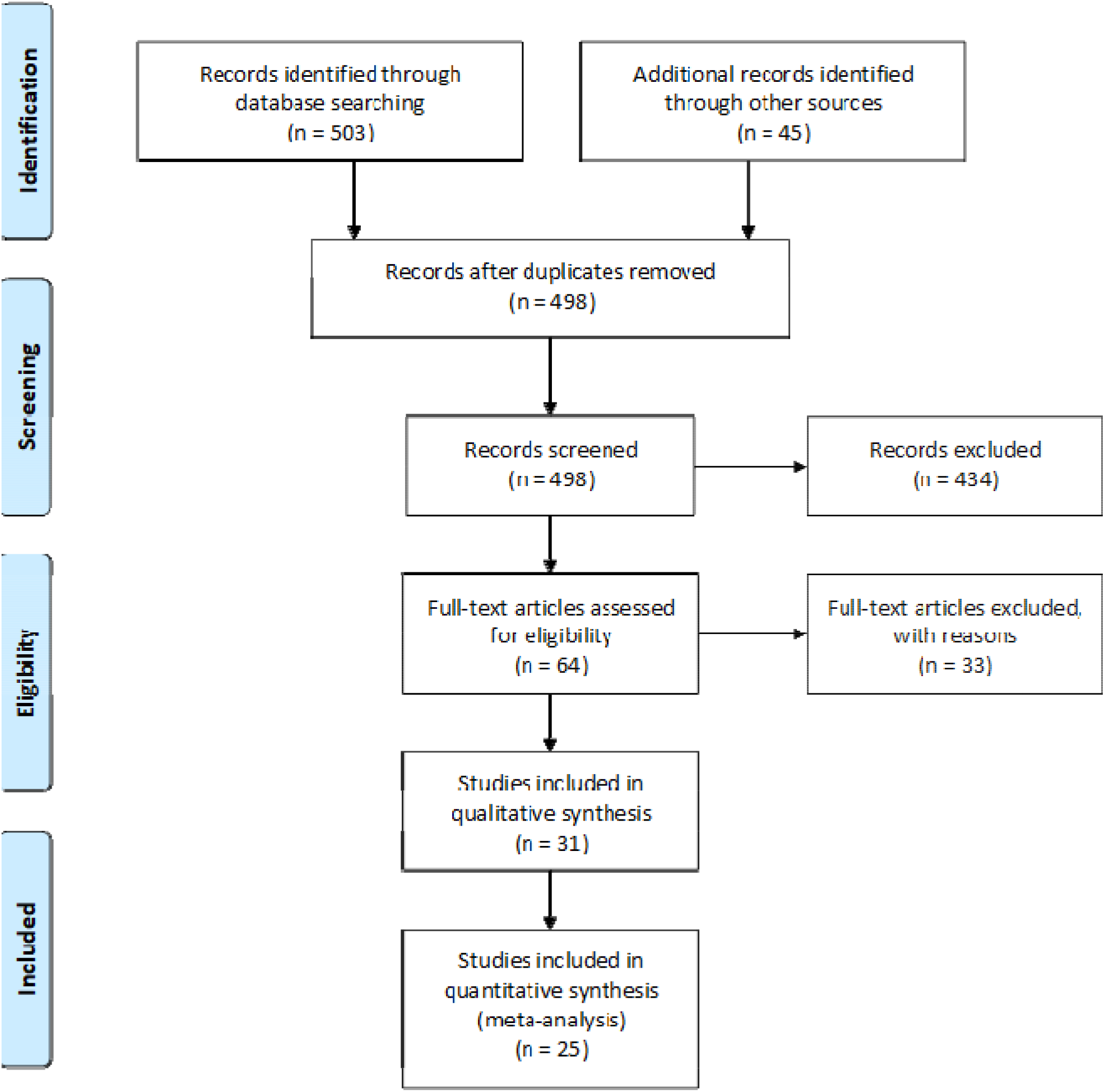
Flow chart of PTB incidence and mortality proportions associated to COVID-19 in Sub-Saharan Africa

The selected articles reported data from South Africa (n = 9), Nigeria (n = 4), Democratic Republic of Congo (n = 2), Angola (n=1), Uganda (n = 1), Zambia (n = 5), Ethiopia (n=2) and Kenya (n = 1) (**Figure 2 and Supplemental material Table 1**)

**Figure 2:**
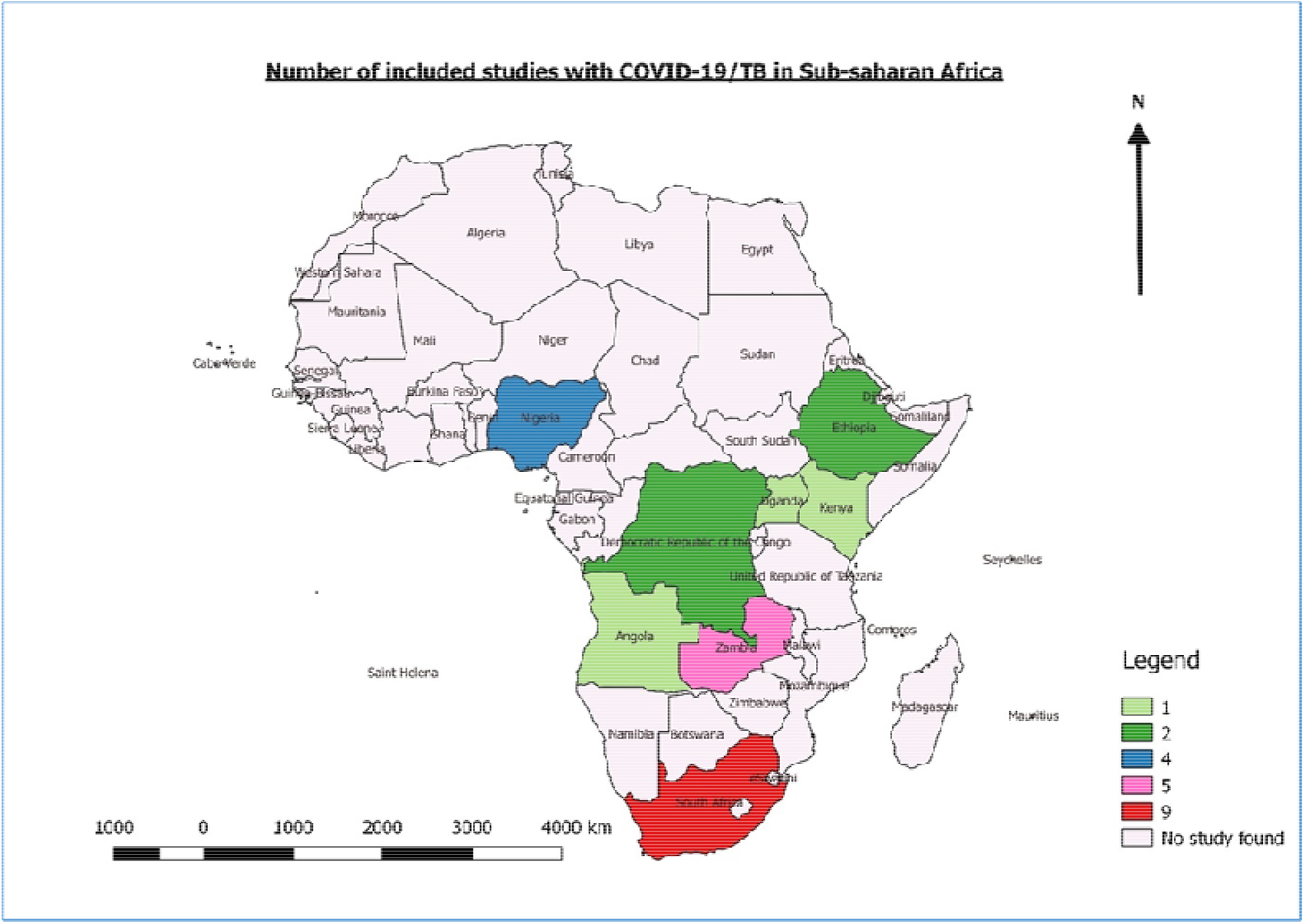
Distribution of included studies in Sub-Saharan African countries

### 2.2. Included studies

Twenty-five studies were included for quantitative analysis. All the studies analysed clinical characteristics and co-morbidities of COVID-19 patients in SSA. Among them, twenty studies included the incidence proportion of COVID-19 associated with PTB [16-35], and nine studies included the case fatality rate of COVID-19 associated with TB [13, 16, 26, 29, 34, 36-39].

The minimum age for the study population was 13 years and the maximum was 72 years. Incidence and case fatality of PTB associated with COVID-19, stratified by sex were not obvious as all the studies included common co-morbidities associated with COVID-19 without sex stratification. The median study duration was 15 months (ranging, from 4 months to 45 years) and the study period ranged from 2020 to 2021. Eleven studies used retrospective case identification [13, 26-33, 36, 39], six prospective case identification [16-20, 35], and three were case series [20, 37, 38], three Cross-sectional studies [22-24], one cross-sectional and retrospective cohort study [34] and one case-control study [25] (**Supplementary material Table 1**).

### 2.3. Study quality

The Newcastle-Ottawa scale (NOS) was used to determine the methodological validity of included research for determining the consistency of cohort, case-control, and descriptive studies in meta-analyses. The three essential components of this strategy are range, comparability, and exposure **(Supplementary material Table 2 and Supplementary material method 3)**. For case-control and cohort studies, the NOS employs a star system with ratings ranging from 0 to 9 **(Supplementary material Table 2 and Supplementary material method 3)**. We considered a study with a higher score than the six of each type of study to be a high-quality study because the criteria for determining whether a study is a high or poor quality are unknown. Two studies scored eight, five scored seven, eight scored six, five scored five, and one scored four. **Supplementary material Table 2** shows the NOS scores for the studies that were included.

### 2.4. Outcomes measurement

In this review, we defined the incidence proportion as the number of new cases of COVID-19 associated with PTB over the total number of people in the population at risk for having COVID-19 during a specified period (**Supplementary material method 3**). The case fatality rate was defined as the total number of new deaths due to COVID-19 associated with TB divided by the total number of COVID-19 associated with PTB. The incidence and case fatality rates were summarized as specific period cases per 100 or 1, 000 (**Supplementary material method 3**). The incidence and case fatality rates of COVID-19 associated with TB were our primary outcomes. The incidence and case fatality rates of COVID-19 associated with previous and current TB were the secondary outcomes.

### 2.5. Meta-analysis and meta-regression

#### 2.5.1. Incidence proportion of COVID-19 associated with TB

In total, 20 studies were identified for the incidence proportion of COVID-19 associated with PTB in SSA. The pooled RR-P [95% CI] was 3% [2%-5%] (**Figure 3**). The test for heterogeneity was statistically significant with (*I*^2^ = 99.76 P<0.0001) (**Figure 3**).

**Figure 3:**
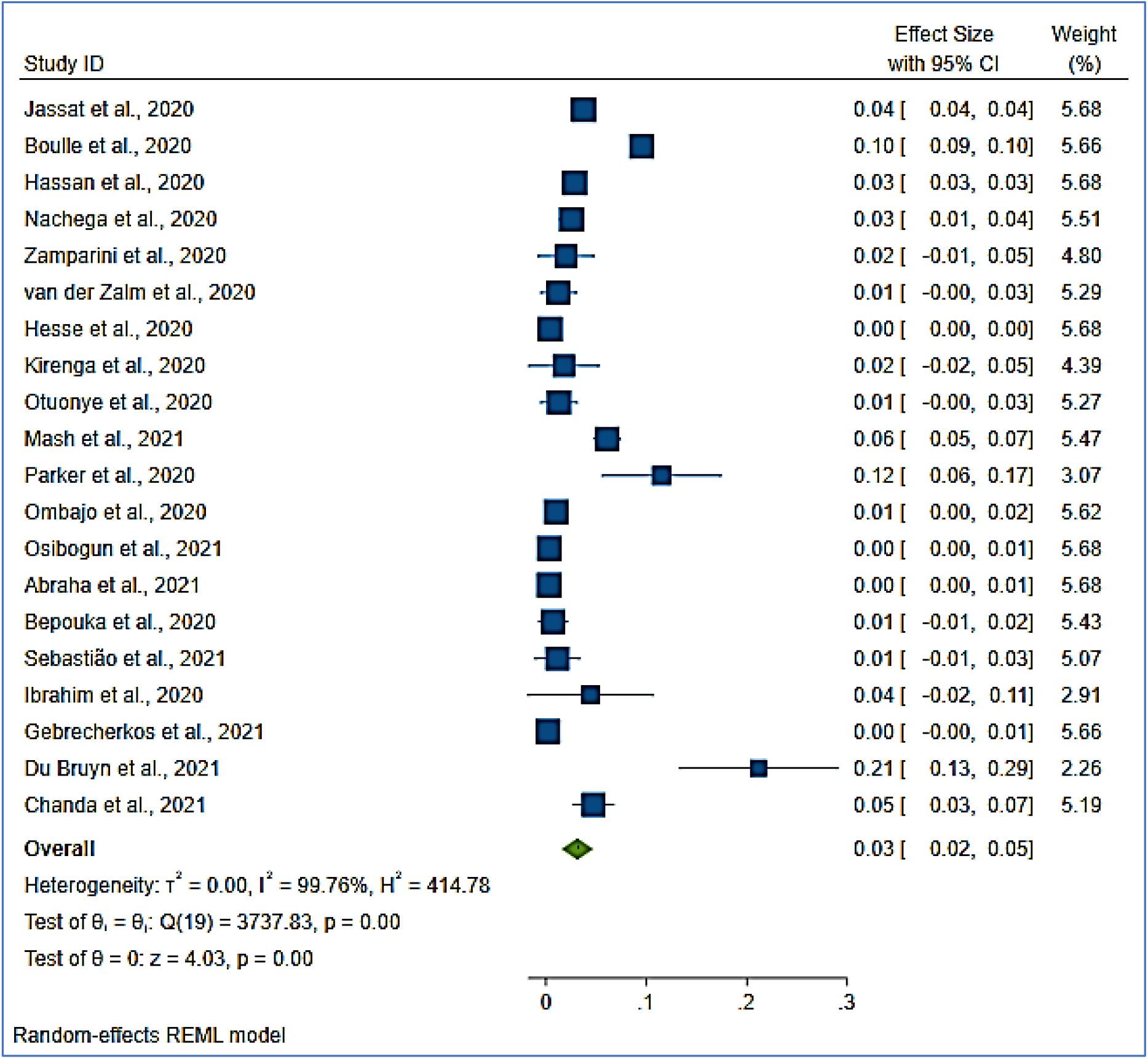
pooled incidence proportion of COVID-19 associated with PTB in sub-Saharan Africa

#### 2.5.2. Incidence proportion of COVID-19 associated with current and previous TB

Only six of the twenty studies included in the pooled result of the incidence proportion rate of COVID-19 associated with TB clearly classified current and previous TB. The pooled current and previous TB incidence proportion rate was 28 [13-43] per 1,000 population (**Figure 4**). Previous TB had the highest incidence proportion rate with 46 [19-73] per 1,000 population (**Figure 4**). The incidence proportion rates for current TB and current and past TB were 14 [6-22] and 10 [9-11] per 1,000, respectively (**Figure 4**).

**Figure 4:**
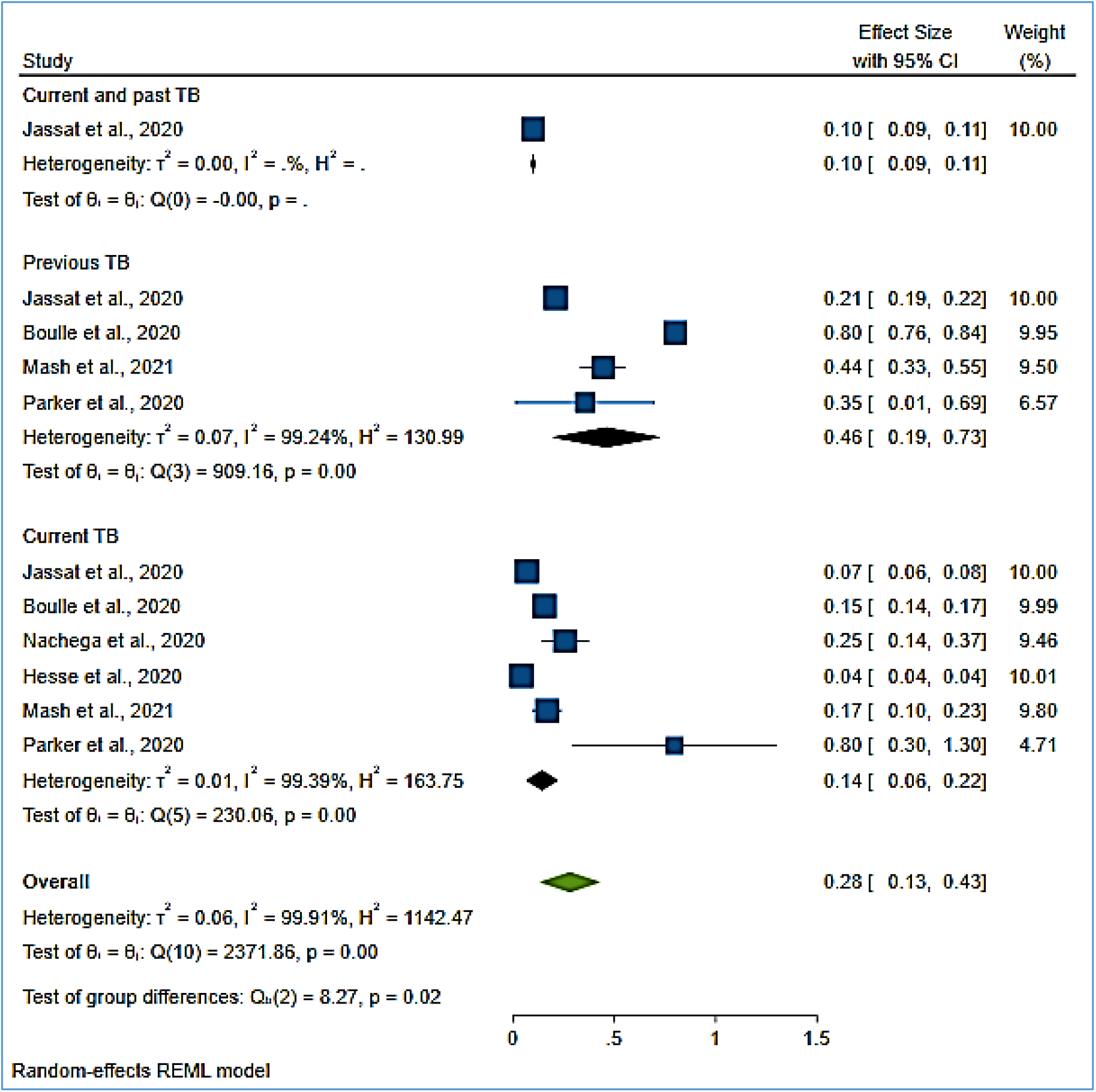
pooled incidence proportion of COVID-19 associated with previous and current PTB.

#### 2.5.3. Case fatality rate of COVID-19 associated with TB

We identified ten studies [13, 16, 17, 26, 29, 34, 36-39] meeting the inclusion criteria relating mortality proportion of COVID-19 associated with PTB in SSA. The pooled RR-P [95%CI] estimates for mortality proportion among patients with COVID-19 associated with TB were 7% [1%-12%] for clinical PTB diagnostic and 25% [3%-47%] for post-mortem PTB diagnostic. The overall pooled RR-P was 13% [4%-23%] (**Figure 5**). Heterogeneity between studies was high (*I*^2^ = 98.82, P <0.001) however, the test for subgroup analysis did not show any difference between the groups (**Figure 5**).

**Figure 5:**
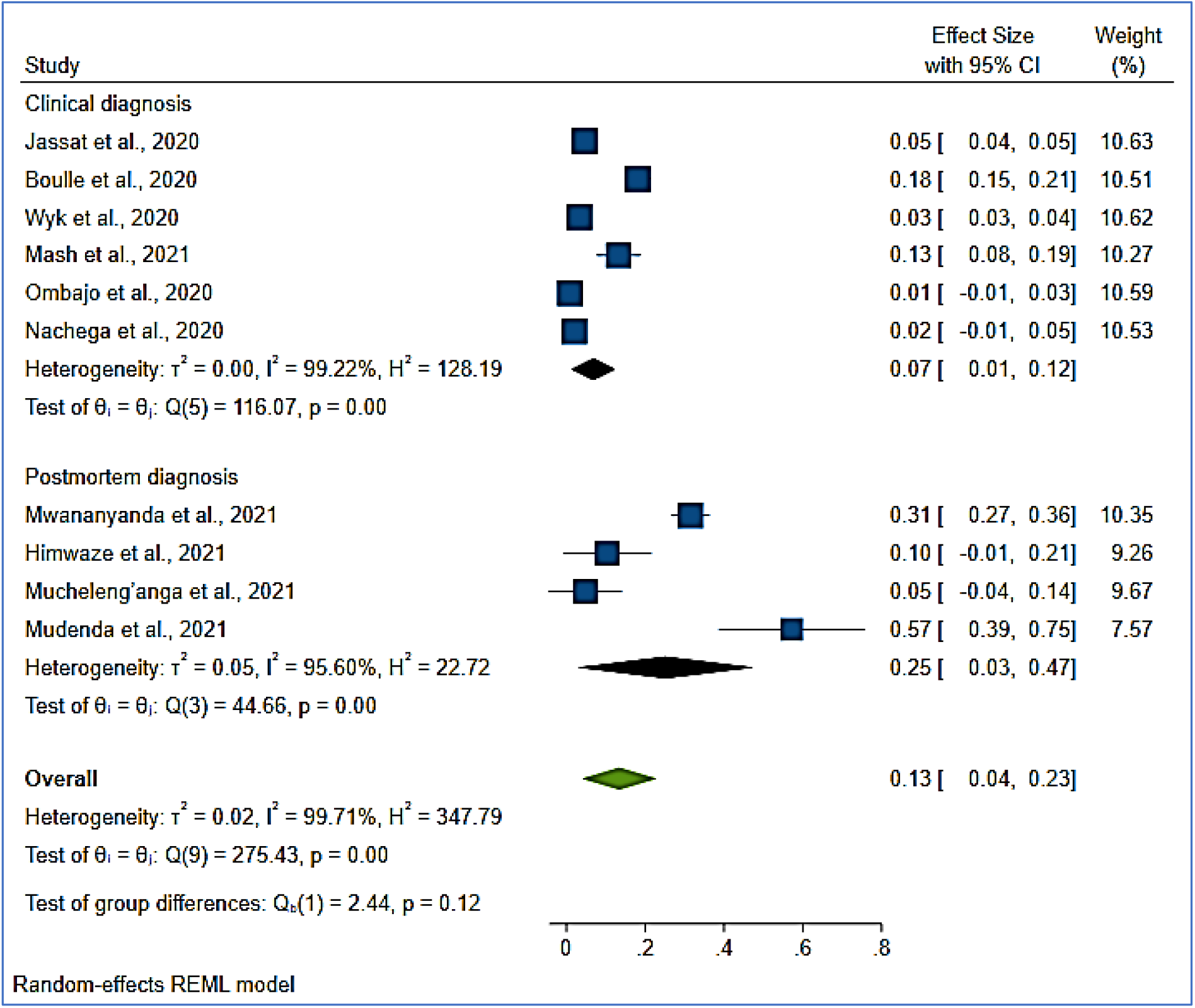
pooled case fatality rate of COVID-19 associated with PTB in sub-Saharan Africa

#### 2.5.4. Case fatality rate of COVID-19 associated with current and previous TB

In the meta-analysis of the case fatality rate of COVID-19 associated with current and previous TB, four studies clearly categorized clinical diagnoses of current and previous TB. Previous TB had the highest case fatality rate of 8% [3%-19%] (**Figure 6**). Current TB and current and past TB had a case fatality of 3% [1%-4%] and 1% [1%-1%], respectively (**Figure 6**).

**Figure 6:**
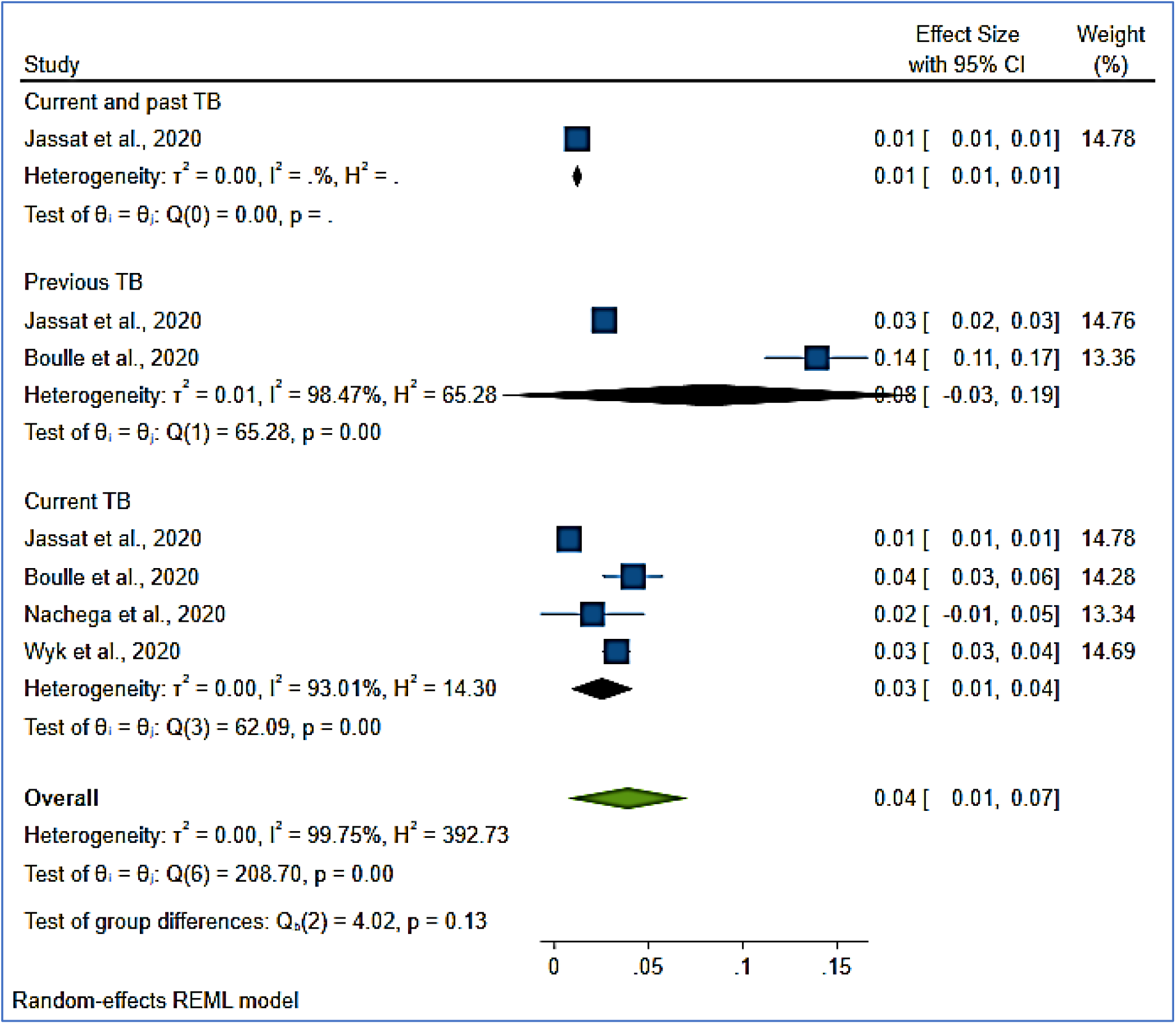
pooled case fatality rate of COVID-19 associated with current and previous PTB

#### 2.5.3. Meta-regression

We built a multivariate meta-regression model including cumulative COVID-19 cases, HIV prevalence, and TB incidence to explore heterogeneity between studies. People living with HIV (all ages), TB incidence rate, and cumulative COVID-19 cases were (7, 800, 000; 360 in thousands; 3, 533, 106) for South Africa, (1, 700, 000; 440 in thousands; 231, 413) for Nigeria, (1 700 000; 59 in thousands, 221, 880) for Zambia, (1, 500, 000; 157 in thousands; 382, 371) for Ethiopia, (510, 000; 278 in thousands; 70, 059) for the Democratic Republic of Congo, (1, 400, 000; 140 in thousands; 270, 899) for Kenya, (340, 000; 112 in thousands; 65, 938) for Angola and (1, 400, 000; 253 in thousands; 130, 178) for Uganda [1, 40, 41]. This model showed that the variability across studies was explained by COVID-19 cumulative cases by countries (P= 0.032), HIV prevalence (P= 0.041), and TB incidence (P= 0.002).

#### 2.5.4. Publication bias

Egger’s and Mazumdar’s rank correlation test and Begg’s funnel plot were used to evaluate publication bias quantitatively and qualitatively respectively. Asymmetry was found in the plot including COVID-19/TB incidence proportion (**Figure 7**). Both Egger’s and Mazumdar’s rank correlation tests did not exhibit obvious publication bias in different studies included in the review because the P-values of both tests for COVID-19/TB incidence rate were (−1.10, P = 0.285) and (−0.26, P = 0.795), respectively. Furthermore, P-values of both tests for COVID-19/TB mortality rate were [Egger’s test (t = 0.49, p = 0.642)] 0.173 and [Begg and Mazumdar’s rank correlation test (z = -0.83, p = 0.404)], respectively.

**Figure 7:**
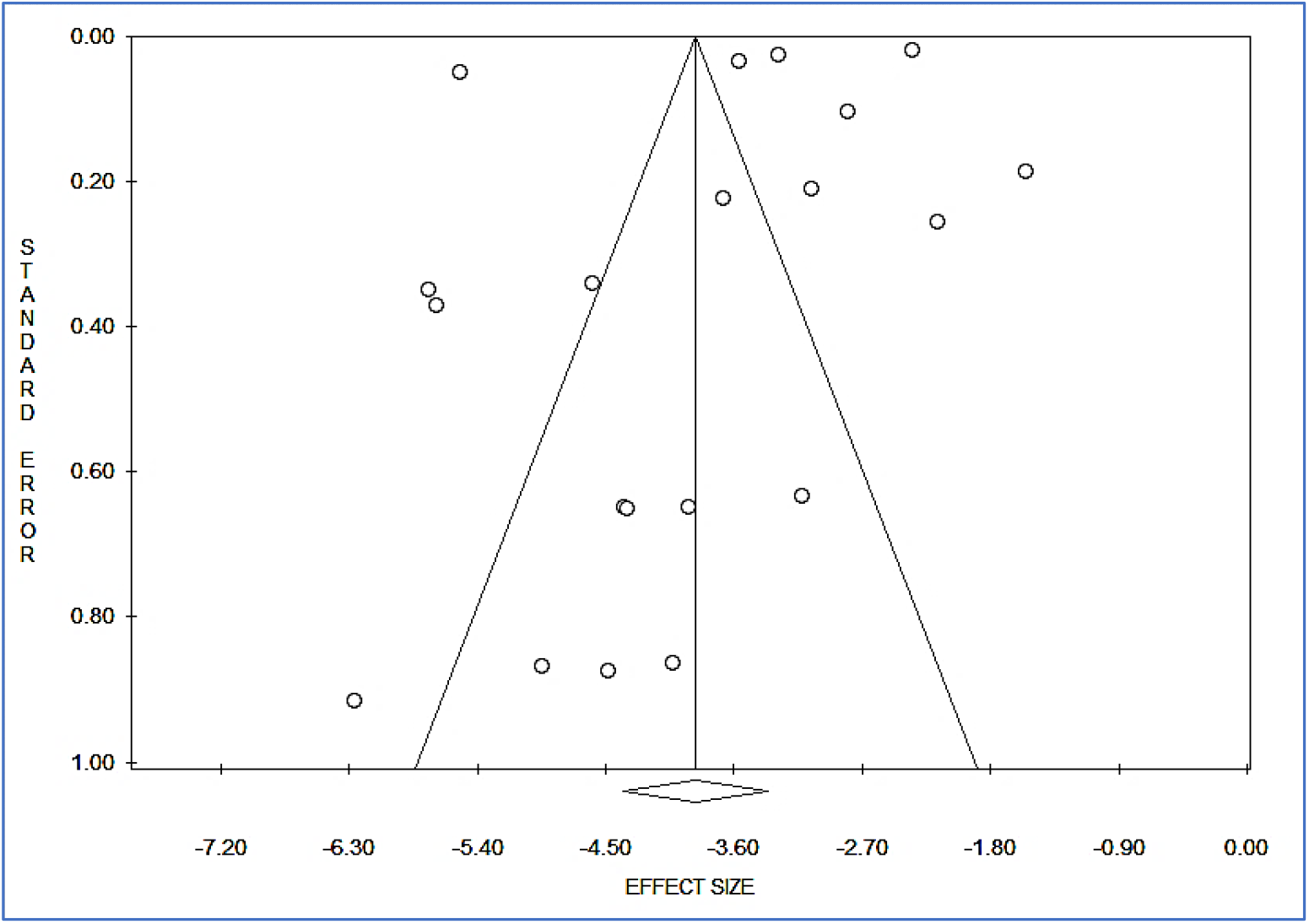
Funnel plot of incidence proportion of COVID-19 associated with PTB in sub-Saharan Africa

#### 2.5.5. Sensitivity analysis

For all the studies including in COVID-19/TB incidence and case fatality rate in SSA, sensitivity analysis was performed by sequential omission of every study respectively Prometa 3 **(Supplementary material method 3)**. For every incidence and mortality rate, the RR-P was not significantly influenced by omitting any single study.

## 3. Discussion

The objective of this review was to help us to estimate the burden of COVID-19 associated with TB in SSA; the meta□analysis including twenty studies and 191, 250 COVID-19 infected cases demonstrated that the overall pooled incidence proportion was 3% [2%-5%]. Our COVID-19/TB incidence was higher than the incidence found in a recent meta-analysis of forty-three studies which showed the pooled estimate for the proportion of active PTB was 1.07% [0.81%-1.36%] [42]. This systematic review included more studies conducted in low and moderate TB prevalence compared to our review which included studies in high-burden TB countries in SSA. Additionally, this review only included active PTB associated with COVID-19, compared to our study which included COVID-19 cases with previous and/or active TB.

The case fatality rate of COVID-19/TB including nine studies and 11,480 COVID-19 deaths was 13% [4%-23%]. However, the meta-analysis of subgroup analysis (clinical vs post-mortem TB diagnostics) showed post-mortem PTB diagnostic counted higher case fatality rate than clinical PTB diagnostic with 25% [3%-47%] for post-mortem TB diagnostic compared to 7% [1%-12%] for PTB clinical diagnostic. Our case fatality rate was higher than a meta-analysis including seventeen studies with 42,321 COVID-19 patients, of whom 632 (1.5%) had TB, reported on deaths due to COVID-19 [42]. Our high case fatality rate may be justified by the same reasons referred to COVID-19/TB incidence. Interestingly, our review has shown that TB was among the most common co-morbidity in COVID-19 patients in SSA. This is consistent with findings in other studies including high TB post-mortem diagnostic in SSA [43, 44]. Referring to a meta-analysis showing that TB exposure was in the high-risk COVID-19 group (OR 1.67, 95% CI 1.06–2.65) [8]. COVID-19/TB clinical diagnostic may be underestimated in SSA as post-mortem TB diagnostic has shown a high mortality rate. Tamuzi et al. have suggested an algorithm for suspected COVID-19/TB diagnostic in high-burden HIV/TB countries [8]. This algorithm may reflect the true COVID-19/TB incidence in high-burden TB countries and reduce COVID-19/TB severity rate OR 4.50 (95% CI 1.12–18.10) and mortality (OR 2.23, 95% CI 1.83–2.74) compared to non-TB group [8].

This deleterious synergism of SARS-CoV-2 and *Mycobacterium tuberculosis* increases the risk of COVID-19-associated morbidity and mortality [8], and patients with PTB may increase the severity of COVID-19 and death due to chronic lung disease and immunosuppression. In fact, advanced PTB is characterized by significant collagen deposition and fibrosis [45-46], although tissue remodelling during fibrosis is a healing process, extensive fibrosis with scar formation impairs lung function [47]. A study reported a series of 454 cases of massive fibrosis with evidence of TB in 40% [48]. Furthermore, Angiotensin-converting enzyme 2 (ACE2) has been reported to play a protective role in lung fibrosis [49]. In lung biopsy specimens of patients with lung fibrosis, ACE2 messenger RNA (mRNA) and enzyme activity decreased significantly [49, 50]. Interestingly, SARS-CoV-2 spike protein decreases the amount of ACE2 expression during viral infection [51]. Decreased ACE2 expression results in increased Angiotensin II (ANG-II) levels and contributes to lung fibrosis and pulmonary failure [49]. In three different acute lung injury models, loss of ACE2 expression precipitated serious acute lung failure, while recombinant human ACE2 (RhACE2) attenuated acute respiratory distress syndrome (ARDS) and further decreased ANG-II levels in the lungs [52, 53]. In addition to transforming growth factor beta (TGF-β), and ACE2, other pathways can contribute to SARS-CoV-2 mediated lung fibrosis. Monocyte chemoattractant protein 1 (MCP-1) is a chemokine that causes lung fibrosis. In addition, there are permanent changes in lung architecture after TB due, in part, to aberrant wound-healing processes [54]. Regulation of the TGF-β signalling pathway was also associated with elevated levels of collagen in lung lesions prior to and during TB [54, 55]. The TGF-β activation pathways in both SARS-CoV-2 and PTB contribute to the production of fibrin, collagen, and secreted proteases (Matrix metalloproteinases) associated with human cavities involved in the formation of fibrosis and tissue remodelling [47]. In summary, the presence of cavitary lesions, fibrosis, and extensive lung pathology was then identified as a major risk factor for poor COVID-19/TB outcomes, which could be explained by reduced drug penetration due to minimal blood supply in fibrotic lung sites [56]. This is consistent with our study findings, which revealed that previous TB had the highest incidence proportion and case fatality rate among COVID-19 when compared to current TB. In fact, previous TB patients may be more prone to cavitary lesions, fibrosis, and extensive lung pathology. Lastly, the transient immunosuppression characterized both COVID-19 and current TB, a reason for poorer IgG antibody response and a delayed viral clearance in co-infected SARS-CoV-2 patients, and the use of corticoid therapy in SARS-CoV-2 added even more on immunosuppression [8]. This could be explained by significant cases of current TB who are likely on TB treatment or were latent TB infection (LTBI) among HIV-infected cases included in the pooled result of the current TB studies by Jassat et al., 2020; Boulle et al., 2020; and Parker et al., 2020 (Supplementary Table 1).

There are several implications to our findings in screening TB concomitantly to COVID-19 in SSA. Our findings indicate that the risks of COVID-19 associated with previous and/or current TB may be underestimated in SSA, as this co-infection is poorly reported. Although there is a paucity of accurate epidemiological data about COVID-19 associated with previous and/or current TB, the fatality rate is estimated high. Therefore, COVID-19 associated with TB should be taken in the context of proper history taking, accurate diagnostic tools, and clear management [8]. Then, clinicians dealing with a possible SARS-CoV-2 patient from a high-burden TB region should never forget TB as a coexisting pathology.

There was substantial variation between studies that looked at the incidence and case fatality rates. This heterogeneity can be explained by a model that includes cumulative COVID-19 cases, HIV prevalence, and TB incidence, all of which vary significantly across countries. Meta-regression revealed statistically significant p-values for the relationship between effect size and our model. Furthermore, the RR-P was not significantly influenced by omitting any single study in a sensitivity analysis performed by sequentially omitting every study for every incidence and mortality rate. To assess publication bias quantitatively and qualitatively, Egger’s and Mazumdar’s rank correlation test and Begg’s funnel plot were used. Asymmetry was found in the plot that included the COVID-19/TB incidence proportion (Fig 7). Because the P-values of both tests for COVID-19/TB incidence rate were (−1.10, P = 0.285) and (−0.26, P = 0.795), respectively, neither Egger’s nor Mazumdar’s rank correlation tests showed obvious publication bias in different studies included in the review. Furthermore, the P-values for [Egger’s test (t = 0.49, p = 0.642)] 0.173 and [Begg and Mazumdar’s rank correlation test (z = -0.83, p = 0.404)] for COVID-19/TB mortality rate were 0.173 and 0.404, respectively.

Our systematic review is limited by the quality of the included studies: first, the majority involved retrospective data analyses, which increase the risk of bias associated with the recording of baseline data, the need for imputation, and potential selection bias. Retrospective studies did not report how missing data were managed or if imputation was used. Considerable risk of selection bias was noted in retrospective and cross-sectional studies. Finally, the results of the case fatality meta□analysis should be interpreted with caution because data pooling the post-mortem PTB diagnostic and previous TB were all extracted in studies conducted in Zambia [13, 37-39] and South Africa [16, 26], respectively. This could limit the external validity of the review. Future studies should be meticulously designed with high□quality and systematic methods of PTB diagnostic associated with COVID-19. This will play a substantial role in reflecting the true incidence and mortality rates of COVID-19/TB in SSA. Furthermore, post-COVID pulmonary fibrosis (PCPF) or long COVID-19-related interstitial lung disease (LC-ILD) associated with current or previous TB requires more attention and research in high HIV/TB burden regions such as SSA.

## 4. Conclusion

This is a systematic review of the incidence proportion and case fatality rate of COVID-19 associated with tuberculosis in SSA. This study found that the incidence of tuberculosis associated with COVID-19 is higher in SSA than in the rest of the world, as are the case fatality rates. Previous TB among COVID-19 patients had the highest incidence proportion and case fatality rates. However, due to the lack of specific COVID-19/TB diagnostic tools, COVID-19 associated with PTB may be underreported in studies conducted in SSA. This is supported by the high case fatality rate of COVID-19/TB in post-mortem diagnostic. Large-scale cohort studies with a sufficiently clear tool on previous and/or current PTB diagnostic tools are required to confirm COVID-19/TB incidence and case fatality.

## Data Availability

All data produced in the present study are available upon reasonable request to the authors

## 5. Abbreviations

ACE2: Angiotensin-Converting Enzyme 2
ARDS: Acute respiratory distress syndrome
COVID-19: Coronavirus disease 2019
HIV: human immunodeficiency virus
LC-ILD: long COVID-19-related interstitial lung disease
MCP-1: Monocyte Chemoattractant Protein-1
mRNA: messenger RNA
NOS: Newcastle-Ottawa scale
PCPF: post-COVID pulmonary fibrosis
PRISMA: Preferred Reporting Items for Systematic Reviews and Meta-Analyses
PTB: pulmonary tuberculosis
SSA: sub-Saharan Africa
SARS-CoV-2: Severe acute respiratory syndrome coronavirus 2
TB: tuberculosis
TGF-β: transforming growth factor
WHO: World Health Organization

## 6. Footnotes

### Contributions

JLT conceived the study and developed the protocol. JLT did the literature search and selected the studies. JLT and GL reviewed the methodological quality of the study and extracted the relevant information. JLT synthesized the data. JLT wrote the first draft of the paper. GL and PB revised successive drafts of the paper. PSN supervised the review steps. All the authors approved its final version. JLT is the guarantor of the study.

## Ethics approval and consent to participate

Not required.

### Consent for publication

Not applicable.

