## Supplemental table 1 for "The incidence and mortality of COVID-19 related TB disease in Sub-Saharan Africa: A systematic review and meta-analysis"

**Supplementary material Table 1: Characteristics of included studies in the incidence and mortality of COVID-19 related TB infection in Sub-Saharan Africa**

| Study ID | Country | Population/Sample size | Study design | Case fatality rate | Incidence rate |
| --- | --- | --- | --- | --- | --- |
| Jassat et al., 2020 | South Africa | 41,845 COVID-19 hospitalized patients including HIV+ and -. Median age: 52 years (IQ: 40-63)  54.4% of patients were females  5 March-11 August 2020 | Retrospective analysis | Total TB (P)=353/7,662  Current and past TB (P): 92/7,662  Previous TB (P): 202/7,662  Current TB (P): 59/7,662 | Total TB (P) = 1325/35,509    Current and past TB (P): 346/35,509  Previous TB (P): 741/35,509  Current TB (P): 238/35,509 |
| Mwananyanda et al., 2021 | Zambia | 372 deceased individuals of COVID-19. Median age: 48 years (IQR 36-72 years)  June and September 2020 | Retrospective analysis | Total TB (P) =117/372 |  |
| Boulle et al., 2020 | South Africa | 1, 2020, 22,308 were diagnosed  with COVID-19 including HIV + and  COVID-19 deceased cases were older than surviving cases (median age [interquartile  range] 63 years [54-71] vs. 37 [30-48])  March 1, 2020 | Population prospective cohort study | Total TB (P)=113/625  Previous TB: 87/625  Current TB: 26/625 | Total TB (P)=2,128/22,308 Previous TB: 1785/22,308 Current TB: 343/22,308 |
| Hassan et al., 2020 | Nigeria | 25,695 confirmed COVID-19 participants.  March-April 2020 | Retrospective analysis |  | Total TB (P)=732/25,694 |
| Nachega et al., 2020 | Democratic Republic of Congo | 766 confirmed COVID-19 participants. The median (IQR): 46 (34–58) years  March 10, 2020–July 31, 2020 | Prospective analysis | Current TB: 2/100 | Current TB (P)=19/745 |
| Wyk et al., 2020 | South Africa | A total of 2,457 COVID-19 deaths Male: 52% and female: 48%). Only 20.3% of the deaths were in individuals aged <50 years, and 29.4% of those who died were aged ≥70 years.  28 March and 3 July 2020 | Retrospective analysis | Current TB (P)= 81/2,457 |  |
| Zamparini et al., 2020 | South Africa | single-centre case series on the first 100 adult patients with reverse-transcriptase polymerase chain reaction (RT-PCR)-confirmed COVID-19 | Single-centre case series |  | Total TB (P)=2/100 |
| van der Zalm et al., 2020 | South Africa | 159 children aged 0-13 years with a laboratory-confirmed SARS-CoV-2 presented to Tygerberg Hospital (TBH).  17 April to 24 July 2020 | Observational cohort study |  | Total TB (P)=2/159 |
| Hesse et al., 2020 | South Africa | 98,335 individuals with positive SARS-CoV-2 PCR test.  The mean age was 42.3±15.0 years.  4-month period in 2020 | Retrospective analysis |  | Current TB (P)=395/98,335 |
| Kirenga et al., 2020 | Uganda | 56 confirmed COVID-19 participants. 67.9% of the patients were male.  The mean age: 34.2 years with an SD of 15.5 years | prospective cohort study |  | Total TB (P)=1/56 |
| Otuonye et al., 2020 | Nigeria | 154 COVID-19 confirmed patients. The mean age (SD) was 46.16(13.701) | Descriptive study |  | Total TB (P) = 2/154 |
| Mash et al., 2021 | South Africa | 1376 COVID-19 positive patients. The mean age of patients was 46.3 years (SD 16.3 years).  March and June 2020 | Descriptive observational cross-sectional Study by means of a retrospective audit of medical records. | Total TB (P) = 20/151 | P=84/1,376  Previous TB (P) = 61/1,376  Current TB (P) = 23/1,376 |
| Parker et al., 2020 | South Africa | 113 patients with confirmed COVID-19/ HIV + and -. The mean (SD) age of patients was 48 (14) years. Females (n=71; 61%) and males (n=45; 39%). | Single-centre descriptive study |  | Total TB (P) =13/113  Previous TB (P) = 4 /113  Current TB (P) = 9/113 |
| Ombajo et al., 2020 | Kenya | 787 confirmed SARS-CoV2. Median age:  43 years. 64% were male.  14th March 2020 and 17th September 2020 | Multi-center cohort study | Total TB (P) =1/107 | Total TB (P)=8/787 |
| Osibogun et al., 2021 | Nigeria | 2075 COVID-19 confirmed participants  The median age of the patients was 40 (IQR=32 - 50) years. The male to female ratio was 2:1.  From 27 February to 6 July 2020 | Retrospective observational study |  | Total TB (P) =7/2,075 |
| Abraha et al., 2021 | Ethiopia | 2617 patients confirmed  COVID-19 positive. The median age of the cohort was 29 (IQR 24–38) years. Most of our study population were male (63.3%) | Retrospective cohort study |  | Total TB (P)=8/2,503 |
| Bepouka et al., 2020 | Democratic republic of Congo | 141 COVID-19 patients admitted at the Kinshasa University Hospital from March 23 to June 15, 2020, were included in the study. their average age was 49.6±16.5 years. 67.4 % were men (sex ratio 2H: 1F) | Retrospective cohort study |  | Total TB (P)=1/141 |
| Sebastião et al., 2021 | Angola | 622 individuals assessed for the SARS-CoV-2 infection between January to September 2020. The age range varied between 1–92 years old, with an average of 32.3±18.7. 244/622 (39.2%) were female and 378/622 (60.8%) were male. | A cross-sectional study |  | Total TB (P)=1/88 |
| Ibrahim et al., 2020 | Nigeria | 45 patients with the diagnosis of COVID-19. Patients were young and male. | Retrospective study |  | Total TB (P)=2/45 |
| Gebrecherkos et al., 2021 | Ethiopia | 515 individuals enrolled  Most of our study population were male (62.5%). The median age of the cohort was 32 (IQR 26–43) years, the majority (60.7%) being in the age range 24 to 44 years. | Prospective observational cohort study |  | Total TB (P)=1/515 |
| Himwaze et al., 2021 | Zambia | 29 whole body autopsies we had performed of COVID-19 inpatient. mean age=44 ± 15.8years; age range=19-82.  17/29 [58.8%] males | Retrospective descriptive study | Total TB (P) = 3/29 |  |
| Mucheleng’anga et al., 2021 | Zambia |  | Retrospective case series | Total TB (P) =1/21 |  |
| Du Bruyn et al., 2021 | South Africa | 104 SARS-CoV-2 RT-PCR positive | single-centre observational case-control study |  | Total TB (P)=22/104 |
| Mudenda et al., 2021 | Zambia | 29 deceased individuals | Retrospective case series | Total TB (P) = 16/28 |  |
| Chanda et al., 2021 | Zambia | 443 patients | Prospective cohort study |  | Total TB (P) =21/443 |
