## Supplemental table 2 for "The incidence and mortality of COVID-19 related TB disease in Sub-Saharan Africa: A systematic review and meta-analysis"

**Supplementary material Table 2: Results of Assessment of Study Quality and Risk of Bias**

| **N** | **Study ID** | **Quality assessment of included studies.** | | | | |
| --- | --- | --- | --- | --- | --- | --- |
|  |  | **Study designs** | **Selection** | **Comparability** | **Outcome/ exposure** | **Overall quality** |
| 1 | Jassat et al., 2020 | Retrospective analysis | ** | ** | *** | 7 |
| 2 | Mwananyanda et al., 2020 | Retrospective analysis | ** | * | *** | 6 |
| 3 | Boulle et al., 2020 | Population prospective cohort study | **** | * | *** | 8 |
| 4 | Hassan et al., 2020 | Retrospective analysis | ** | * | *** | 6 |
| 5 | Nachega et al., 2020 | Prospective analysis | *** | * | *** | 7 |
| 6 | Wyk et al., 2020 | Retrospective analysis | ** | * | *** | 6 |
| 7 | Zamparini et al., 2020 | Single-centre case series | ** | * | ** | 5 |
| 8 | Van Der Zalm et al., 2020 | Observational cohort study | ** | * | *** | 6 |
| 9 | Hesse et al., 2020 | Retrospective analysis | *** | * | ** | 6 |
| 10 | Kirenga et al., 2020 | prospective cohort study | *** | ** | ** | 7 |
| 11 | Otuonye et al., 2020 | Descriptive study | ** | * | *** | 6 |
| 12 | Mash et al., 2020 | Descriptive observational cross-sectional Study and retrospective | *** | * | *** | 7 |
| 13 | Parker et al., 2020 | Single-centre descriptive study | ** | * | ** | 5 |
| 14 | Ombajo et al., 2020 | Multi-center cohort study | *** | * | *** | 7 |
| 15 | Osibogun et al., 2021 | Retrospective observational study | ** | * | *** | 6 |
| 16 | Abraha et al., 2021 | Retrospective cohort study | ** | * | *** | 6 |
| 17 | Bepouka et al., 2020 | Retrospective cohort study | ** | * | ** | 5 |
| 18 | Sebastião et al., 2021 | A cross-sectional study | *** | * | ** | 6 |
| 19 | Ibrahim et al., 2020 | Retrospective study | ** | * | ** | 5 |
| 20 | du Bruyn et al., 2021 | single-centre observational case-control study | ** | * | ** | 5 |
| 21 | Chanda et al., 2021 | Prospective cohort study | **** | * | *** | 8 |
| 22 | Mudenda et al., 2021 | Retrospective case series | ** | * | ** | 5 |
| 23 | Mucheleng'anga et al., 2021 | Retrospective case series | * | * | ** | 4 |
| 24 | Himwaze et al., 2021 | Retrospective descriptive study | ** | * | ** | 5 |
| 25 | Gebrecherkos et al., 2021 | Prospective observational cohort study | ** | * | *** | 6 |

Newcastle-Ottawa Scale was obtained to assess the selection, comparability, and exposure of the case-control study, while the selection, comparability, and outcome for the cohort study. -: no point; *: one point; **: two points; ***: three points; ****: four points.
