## Supplemental Methods for "The incidence and mortality of COVID-19 related TB disease in Sub-Saharan Africa: A systematic review and meta-analysis"

**Supplementary material 3: Methods**

The review followed a predesigned protocol registered in PROSPERO (CRD42021233387). The systematic review met the criteria outlined in the Preferred Reporting Items for Systematic Reviews and Meta-Analyses (PRISMA) guidelines [1].

1.1. **Search strategy and eligibly criteria**

A search of the literature was systematically conducted using PubMed, Medline, Google Scholar, Medrxix, and COVID-19 Global literature on coronavirus disease. All searches were limited to articles written in English given that such language restriction does not alter the outcome of systematic reviews and meta-analyses. The search was restricted to studies related to the incidence and case fatality rates of COVID-19 related to TB in SSA since December 2019 to November 2021 including the key words and term as follows: “Covid-19 or 2019-nCoV or coronavirus disease 2019 or Novel coronavirus or SARS-CoV-2 ” and “tuberculosis or PTB or TB or Mycobacterium tuberculosis infection” and “mortality rate or death rate or case fatality rate” and “incidence or Incidence proportions or Incidence rate or incidence rate or attack rate” and “Angola or Benin or Botswana or Burkina Faso or Burundi or Cameroon or Cape Verde or "Central African republic" or Chad or Congo or "Democratic Republic of Congo" or DRC or Djibouti or Equatorial guinea or Eritrea or Ethiopia or Gabon or Gambia or Ghana or Guinea or Bissau or Ivory coast or "“Cote d’ ivoire" or Kenya or Lesotho or Liberia or Madagascar or Malawi or Mali or Mayotte or Mozambique or Namibia or Niger or Nigeria or Principe or Reunion or Rwanda or "Sao Tome" or Senegal or "Sierra Leone" or Somalia or "South Africa" or Swaziland or Tanzania or Togo or Uganda or Zambia or Zimbabwe or "Central Africa" or "Sub-Saharan Africa" or "East Africa" or "Southern Africa" or “South Africa”, without language restrictions to identify citations from prior to January 2020. The review included observational studies conducted in Sub-Saharan Africa, including the incidence and mortality related to COVID-19 and TB. We included confirmed COVID-19 participants with TB diagnosed previously, currently, or in the post-mortem.

1.2. **Study quality and risk of bias assessment**

The methodological quality of the included studies was independently assessed by two of the authors (JLT and GL). Any inconsistencies were resolved by consensus, and if no agreement was reached yet again, the case was resolved by seeking the views of a third author (PB).

The Newcastle-Ottawa scale (NOS) [2] was used by two reviewers (JTL and GL) to independently assess study quality. The NOS evaluated the case series, cross-sectional, case-control study's selection, comparability, and exposure, as well as the cohort study's selection, comparability, and outcome. The sample with more than 6 stars was of reasonably high quality, and the sample with nine stars reflects the highest ranking. Any discrepancies in the content of the included studies were resolved with the help of another reviewer (PB).

1.3. **Data extraction**

Three levels of screening were performed. The first and second rounds of screening were based on titles and abstracts only while the third round consisted of a review of full-text articles. The first screening was performed by JTL and excluded references that did not contain information on the pathogens of interest or those that were not the study designs of interest (included observational studies only). The second screening was performed independently by JLT and GL with differences solved by consensus. The third level of screening identified those publications related to COVID-19 incidence and mortality associated with PTB in SSA and data extraction was performed on those that met the criteria.

Screening and data extraction were performed by JLT and GL independently reviewing each full-text article. Conflicts were resolved via discussion to achieve consensus, with any remaining disagreements resolved by a third reviewer. Included studies were observational studies that included COVID-19 incidence and/or mortality related to PTB in SSA.

Data were extracted based on the study year of publication, first author's name, country of study, design, setting, target population, sampling method, sample size, total study period, items related to the quality assessment of the study (the score of each item and the overall study quality score), incidence data and mortality of COVID-19 associated with PTB (including attack rates, death rate, clinical and post-mortem COVID-19/TB diagnostic mortality, cumulative incidence and incidence rate, based on the measured 95% CIs and P-values).

1.4. **Statistical analysis**

The main outcomes were the incidence of COVID-19 associated with current or/and previous PTB and the case fatality rates associated with this proportion. This was calculated as the number of persons developing COVID-19 associated with PTB divided by the total number of COVID-19 cases. Standard errors and confidence intervals for a single proportion were derived. P was the proportion of COVID-19/TB infections (previous and/or current TB) or clinical and post-mortem diagnostics of COVID-19/TB deaths and N was the total number of cases of COVID-19 for the incidence rate and total COVID-19 deaths for the case fatality rate.

The pooled COVID-19 incidence proportion and case fatality rate PTB-related were calculated in this meta-analysis. The combination method was based on methodological similarities in the included random effect model studies using Stata version 16 and Prometa 3 software [3]. Forest plots were plotted for all studies to show the separate and pooled incidence and fatality rates and the corresponding 95% CIs. Both COVID-19/TB incidence proportion and fatality rate were measured in terms of the ratios of proportions (RR-Ps). Heterogeneity assessment with the Q-Statistic Test and *I*^2^-Statistics and their corresponding 95% CIs were used to assess the statistical heterogeneity of incidence and mortality in the included studies. The following references were used as the basis for determining the degree of heterogeneity: (1) Heterogeneity values of 0% – 40% will be considered 'maybe not important;' (2) Heterogeneity values of 30% – 60% as 'moderate heterogeneity;' (3) Heterogeneity values of 50% – 90% as 'substantial heterogeneity;' and (4) Heterogeneity values of 75% – 100% as 'significant heterogeneity [19]. The statistical significance level was set at p<0.05 for the Q-test [4]. The subgroup, meta-regression, and sensitivity analysis were used to explore potential sources of heterogeneity if the *I*^2^ value was higher than 75%. We also undertook meta-regression to find out the source of heterogeneity. Lastly, we explored the publication bias with the funnel plot, Egger’s and Begg, and Mazumdar’s rank correlation tests.
